# Families Experience of a Relatives Brain Stem Death Diagnosis: A Systematic Review

**DOI:** 10.1101/2023.03.09.23287057

**Authors:** Ella Cade-Smith, Liam Mackay, Dhuleep Sanjay Wijayatilake, Marc Kingsley, Madiha Shaikh

## Abstract

**AIM:** Being diagnosed as Brain Stem Dead is a very challenging experience for families. Most research regarding brain stem death focuses on Organ Donation and there is currently little research into families’ experience of brain stem death. The aim is to review the family’s experience of brain stem death.

**DESIGN:** Systematic review

**METHOD:** A narrative synthesis was conducted for 9 studies including qualitative and quantitative study designs. Four electronic databases: AHMED (Allied and Complimentary Medicine), Emcare (1995-present), Medline (Ovid) and APA Psych Info (Ovid) were searched. No limit was placed on date of publishing due to this being a relatively under researched topic. The original search was conducted on 4^th^ November 2021 and rerun on 6^th^ December 2022 to ensure the inclusion of any new published studies.

**RESULTS:** Six main themes were identified, including: The Unexpected Prognosis; Coming to terms with brain stem death- grieving process; Request for organ donation; Observing brain stem death testing; The impact of staff on families’ experience; and the lasting impact.

**CONCLUSION:** Families of patients with brain stem death are often left with a lack of understanding surrounding the diagnosis, the process, and the short and long term distress it can cause. There is need for research into family’s experiences and brain stem death testing specifically so that more reliable data can be produced. There is a need to establish national, or international practice surrounding family care in intensive care in cases of brain stem death. This review highlights the importance of establishing specific brain stem death protocols, enabling more effective and consistent support for families.

## Introduction

Despite the first definition of brain stem death (BSD) being identified in 1968 by the Harvard Medical School, there still is not a universally shared concept or recognition accepted (1, 2). General understanding of BSD is the permanent and total loss of all brain function in the brainstem and cerebrum (3) when oxygen or blood supply to the brain is stopped. BSD differs from a vegetative state or coma as with BSD, mechanical ventilation is required for the patient to breathe and is a permanent state with no possibility of recovery.

### Global Differences in BSD Practice

As a result of the lack of universal standard of method for BSD diagnosis, the practise surrounding brain death can vary across countries (4, 5, 6, 7). This suggests that current guidelines need to be re-visited (8). Moreover, some under-developed countries do not have any protocols surrounding BSD (5). Prior to testing for BSD, proof is required to confirm that there is irreversible structural brain damage, and that all possibility of a reversible cause of coma can be excluded. Some components of the formal neurological examination and diagnostic criteria that follows are: apnoea testing, absence of corneal reflex, absence of vestibulo-ocular reflex, demonstration of no motor response to pain, no pupillary response to light and no gag reflex tests (9). These are designed to test the lack of automatic responses, demonstrating permanent damage to the brainstem. Countries will utilize not only different tests, but they also require these tests to be conducted by different professionals with varying expertise (4). Therefore, the differences in perception and practices surrounding brain death globally are substantial, making the concept of BSD complex for both professionals and patients and their families.

### Spiritual Considerations

The notion of brain death is also complex because of individual, spiritual and religious perspectives on what constitutes death. Randhawa 10) noted that in the UK, brain death has ‘in some faith groups, led to considerable debate and remains contested by faith leaders. For example, Popal Popal (11) found that US Islamic physicians with higher religiosity were less likely to equate BSD with cardiopulmonary criteria for assessing death, as well as less likely to consider BSD to signify the soul departing. Also, the Orthodox rabbinic community is divided on whether BSD equates death, with some leading orthodox rabbinic leaders believing that only cardiopulmonary death indicates death (12). This highlights the conflicting nature of the prognosis. Discussions surrounding the validity of BSD have stemmed from these religious, spiritual and moral controversies, as well as further development of BSD criteria within the scientific community (13). These impact understanding of BSD and perception of organ donation. Religions may have differing opinions on the specific organs that can and cannot be donated (14), resulting in a wealth of research over recent years debating BSD and the ethics of organ donation specifically.

### Organ Donation

Specialist organ donation nurses speak with patients’ families about organ donation during the process of a family members’ diagnosis of BSD as donor’s death by neurological criteria increases the likelihood of a successful transplant (15). In some countries, such as Scotland (16) and England (17) donation is presumed, thereby working within an ‘opt-out’ system, whereas other counties operate an ‘opt-in’ system. Furthermore, in some countries like the Netherlands, organ donation is requested before the confirmation of BSD (18). Conversations of this nature are very sensitive for people considering the sudden nature of BSD diagnosis as well as having potential disagreements regarding what the family member and the family themselves would prefer. Consequently, research regarding the family members’ perspective of a relatives BSD is primarily about the organ donation decision and process. This has resulted in less focus on the families’ overall experience of their relatives BSD diagnosis itself.

### The current review

Death by neurological criteria is often an unexpected, tragic, and difficult experience. Patients who are diagnosed as BSD can still appear to be breathing, retaining their usual colour and body temperature, because of the ventilator oxygenating their bodies. That being so, scepticism or confusion of BSD is understandable (19).

In the same way diagnostic criteria is not standardised internationally across Intensive Care Units (ICUs) (20), treatment for families going through this difficult situation is not either. Often communication and language are confused, and this inconsistency can lead to further distress (21). For these reasons, adequate and appropriate care is paramount for the families of those diagnosed as BSD.

Given the lack of research regarding family’s experiences of BSD, a review of the current studies is important to analyse and evaluate peoples’ perceptions and experiences of BSD and the care they received to help understand what best practice looks like, what needs to be improved, and what could be developed to ensure support for families.

## Methods

The review was prospectively registered on PROSPERO (CRD42022332267). Since initial registration the analysis approach and choice of quality bias tool have been altered. Four electronic databases: AHMED (Allied and Complimentary Medicine), Emcare (1995-present), Medline (Ovid) and APA Psych Info (Ovid) were searched. No limit was placed on date of publishing due to this being a relatively under researched topic.

Search terms were created using the PICO framework, resulting in the three concepts: Brain Stem Death, Family and Experience. The following terms were used in our searches: ((brain adj dea*) OR (brain adj stem adj dea*) OR (death adj by adj neurological adj criteria)) AND (family OR families OR (loved adj one*) OR carer* OR partner* OR spouse OR parent* OR guardian* OR sibling* OR brother* OR sister*) AND (experience* OR presence OR attitude* OR acceptance OR opinion* OR perspective*). The original search was conducted on 4^th^ November 2021 and rerun on 6^th^ December 2022 to ensure the inclusion of any new published studies.

Qualitative, quantitative, and mixed methods studies that reported families’ experience of a relatives BSD were included. Studies were included if participants were related to a person with BSD, as a result of blood connection (e.g., sibling, grandmother), marriage (e.g., partner, step-father) or had any significant relationship with the person with BSD.

Studies that were not in English, reviews and individual case reports were excluded.

Title and abstract and full text screening were conducted by ECS. LM independently assessed eligibility of 100% of papers at the full text stage. Any disagreements were resolved with author MS. Author ECS extracted study data using a predefined Microsoft excel form. Extracted data included author(s), year, country, study objective, participants age and relationship to the person with BSD, and families’ experiences of BSD.

A narrative synthesis was used due to the inclusion of various study designs (23). Data was translated using thematic analysis using an inductive approach (23). In this context a process of ‘translation’ of primary themes or concepts reported across studies was used to explore similarities and/or differences between different studies (24). This provided a means of organising and summarising the findings from a range of studies and reflect directly, the main ideas and conclusions across studies, rather than developing new knowledge. Thus, the key qualitative and quantitative findings are presented as they are reported in the original paper. The Joanna Briggs Institute Critical Appraisal Tool (JBI) was used to review the potential risk of bias and quality of publication of qualitative, quantitative, and randomised control trial studies (10 item scale and 13 item scale) and the Mixed Methods Appraisal Tool (MMAT) was used to review mixed method studies (7 item scale). Papers were independently assessed by two authors and any disagreements were resolved through discussion or further evaluation with a third author.

The questions could be answered as ‘yes’, ‘no’ or ‘unclear’. Risk of bias was determined using the following criteria: Calculated as low risk of bias if at least 70% of answers were scored ‘yes’, a moderate risk of bias if 50-69% were scored ‘yes’ and a high risk of bias was considered if there were less than 50% scored ‘yes’ for the individual study (25).

## Results

As shown in the PRISMA flow diagram, a total of 16 papers were assessed for eligibility and 9 met inclusion criteria for this review (Figure 1). Across the 9 studies, 606 family members were included. The studies were from Sweden (*n* = 2), England (*n* = 2), the USA (*n* = 2), the Netherlands (*n* = 1), France (*n* = 1), and Switzerland (*n* = 1). Most studies used a qualitative method (*n* = 6), followed by mixed methods (*n* = 2) and one Randomised Controlled Trial (*n* = 1). The key study details and results are shown in Table 1.

**Figure 1.**
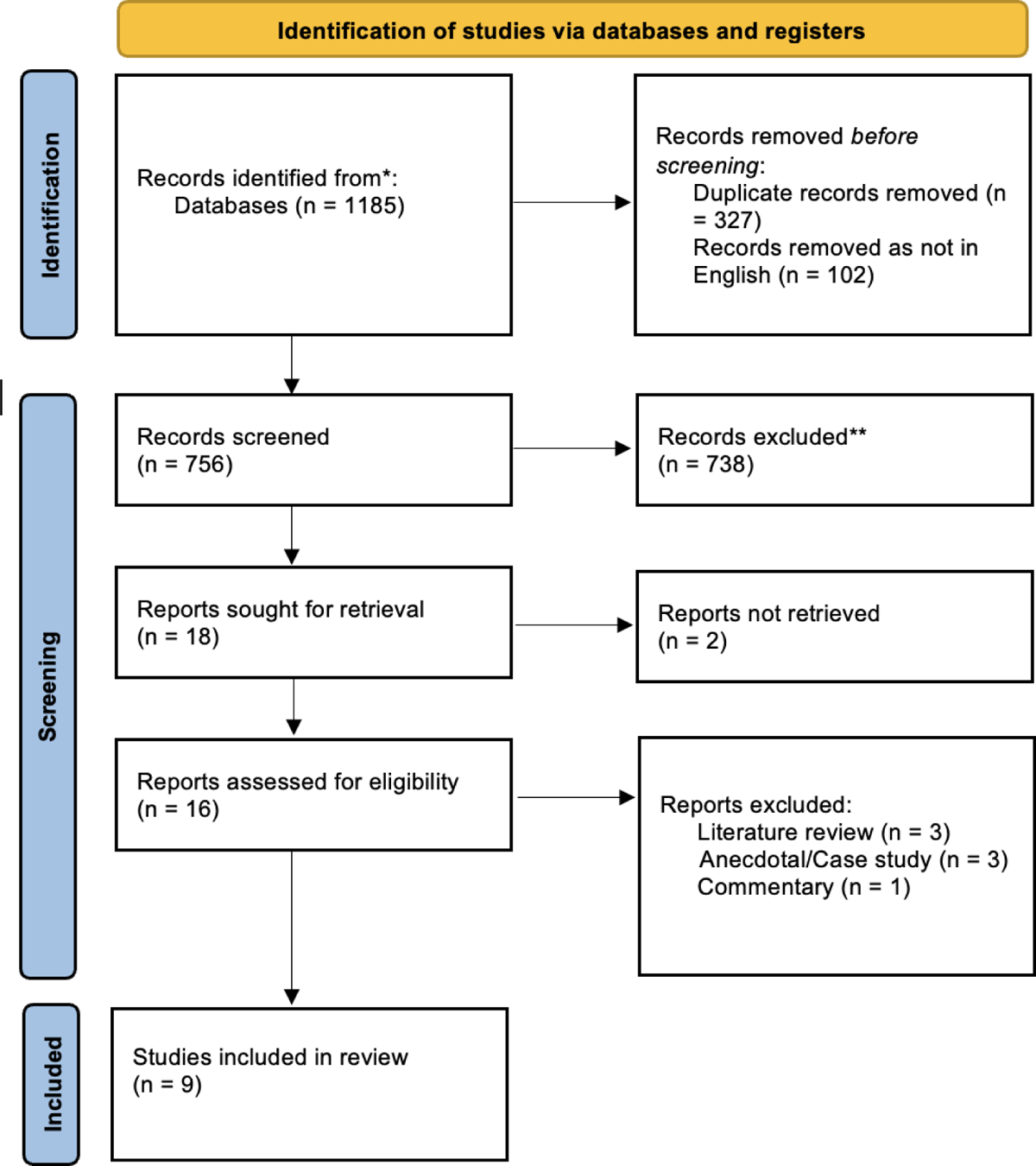
PRISMA Flow Diagram Figure 1. PRISMA flow diagram (22)

**Table 1.**
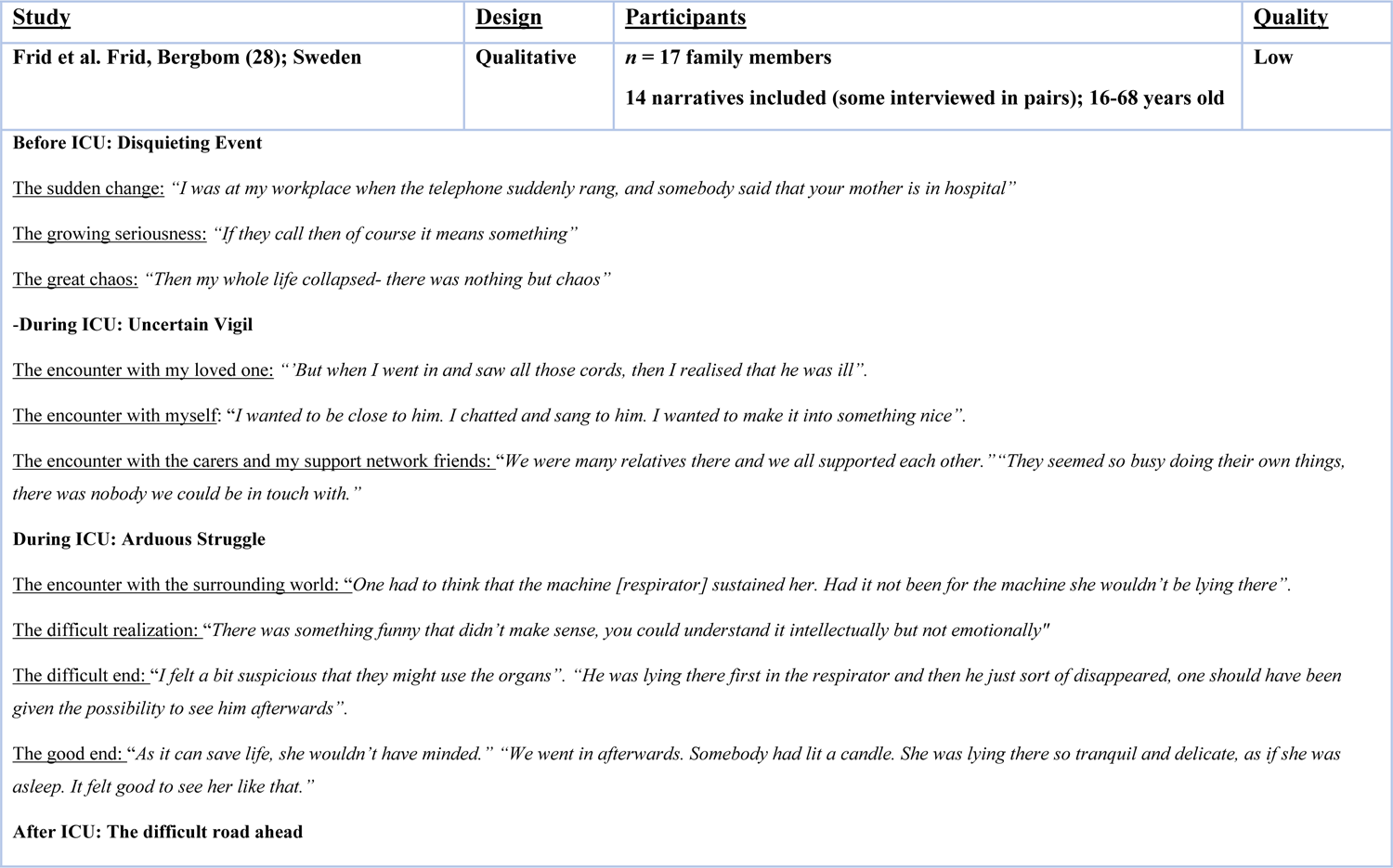
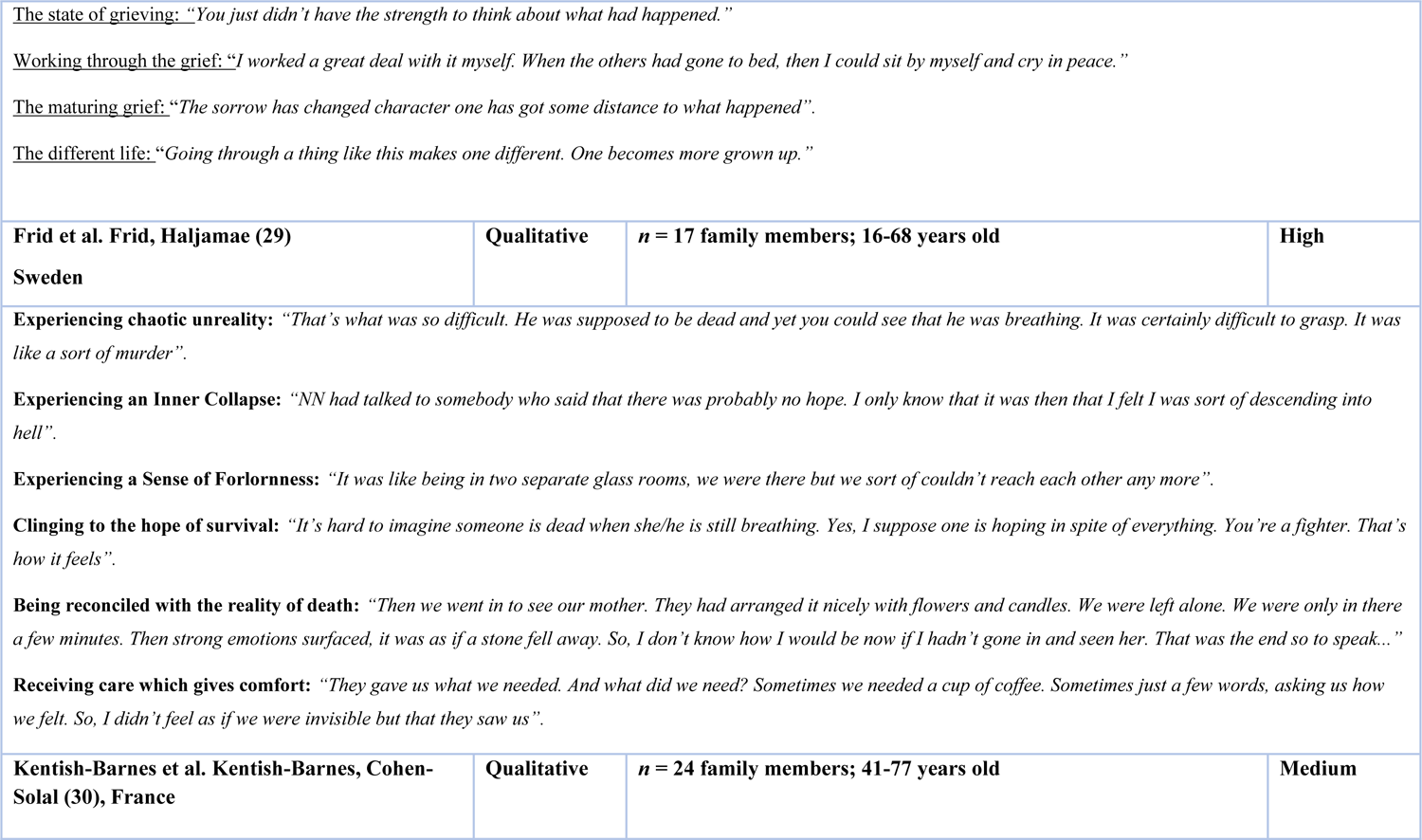
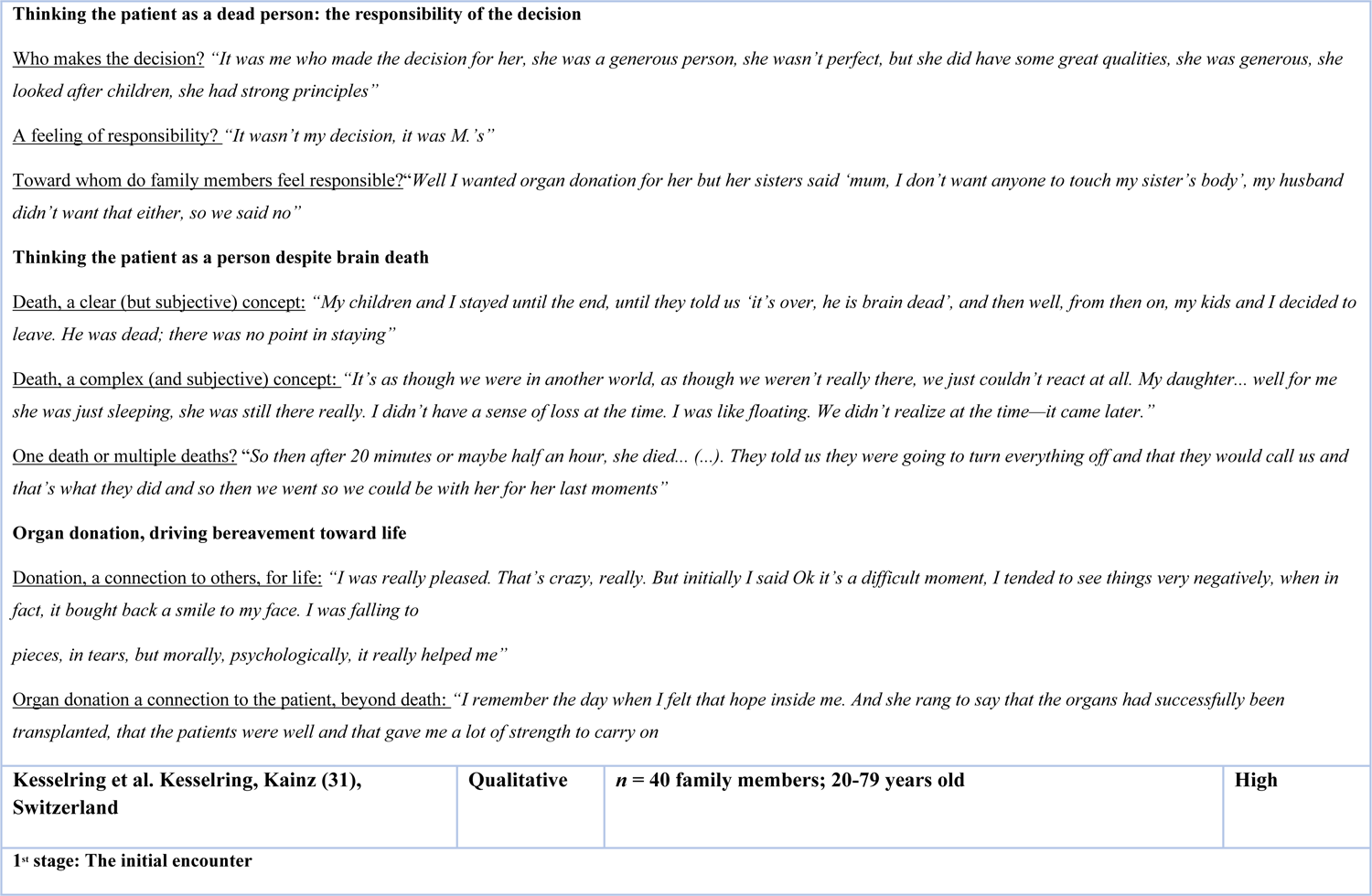
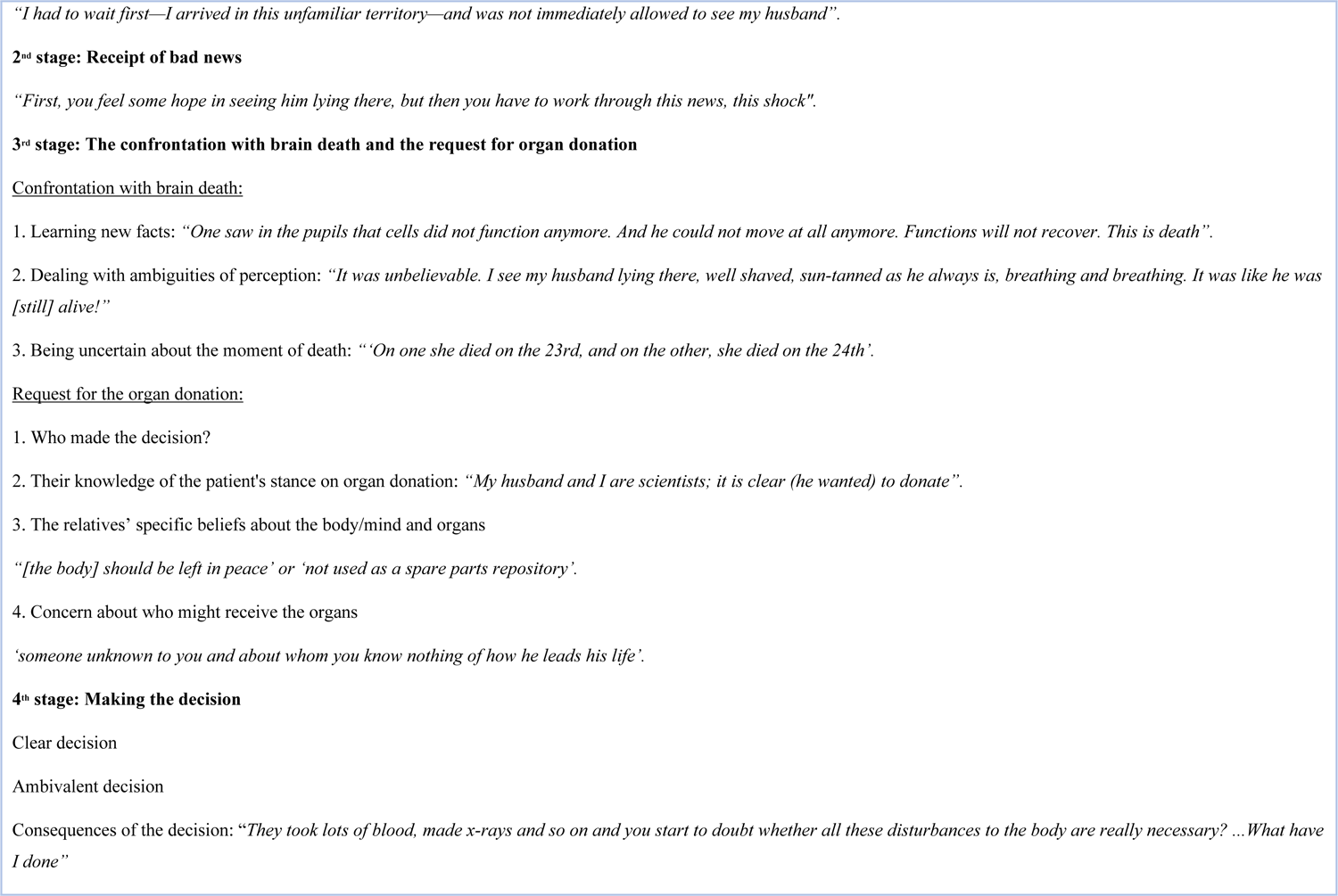
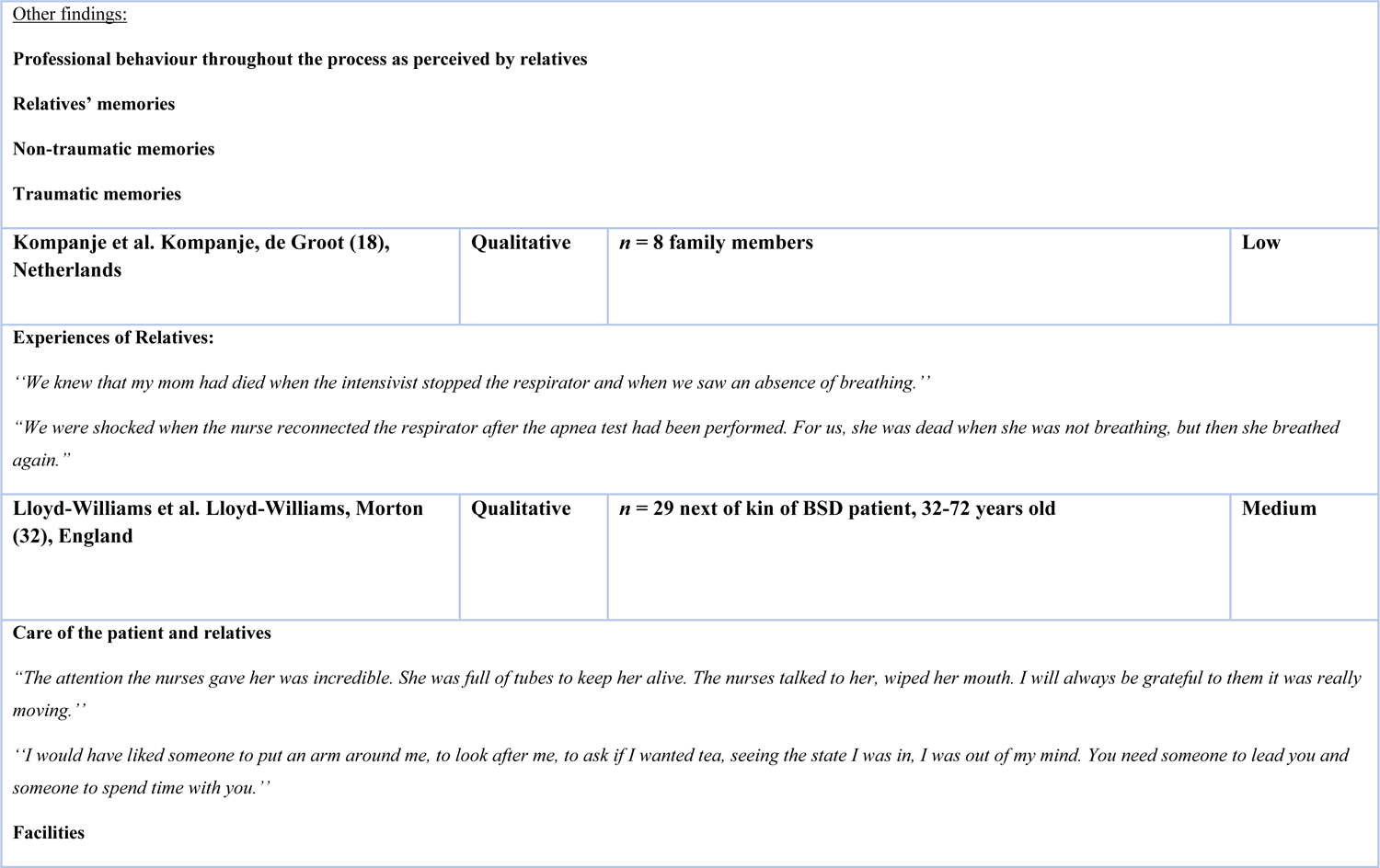
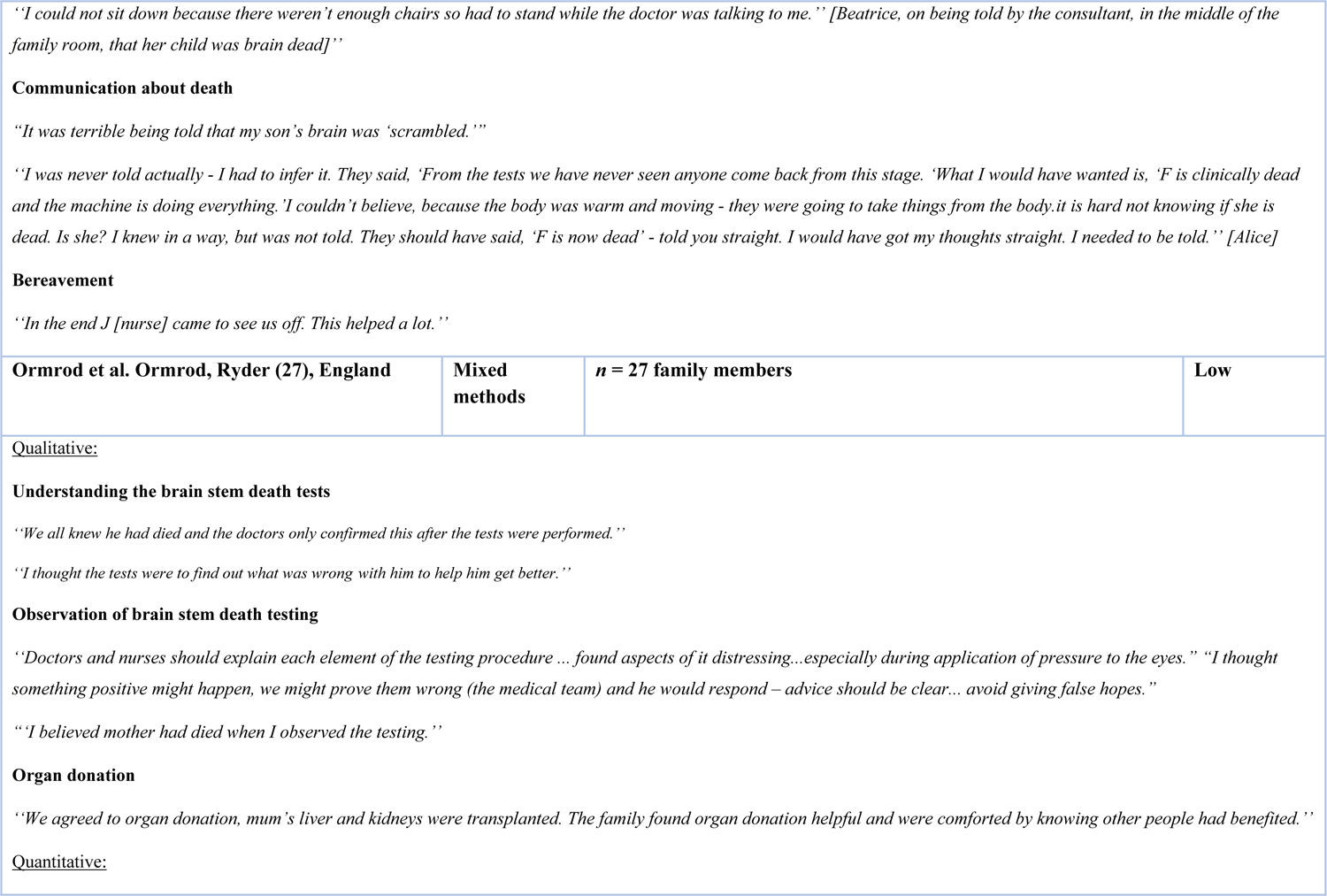
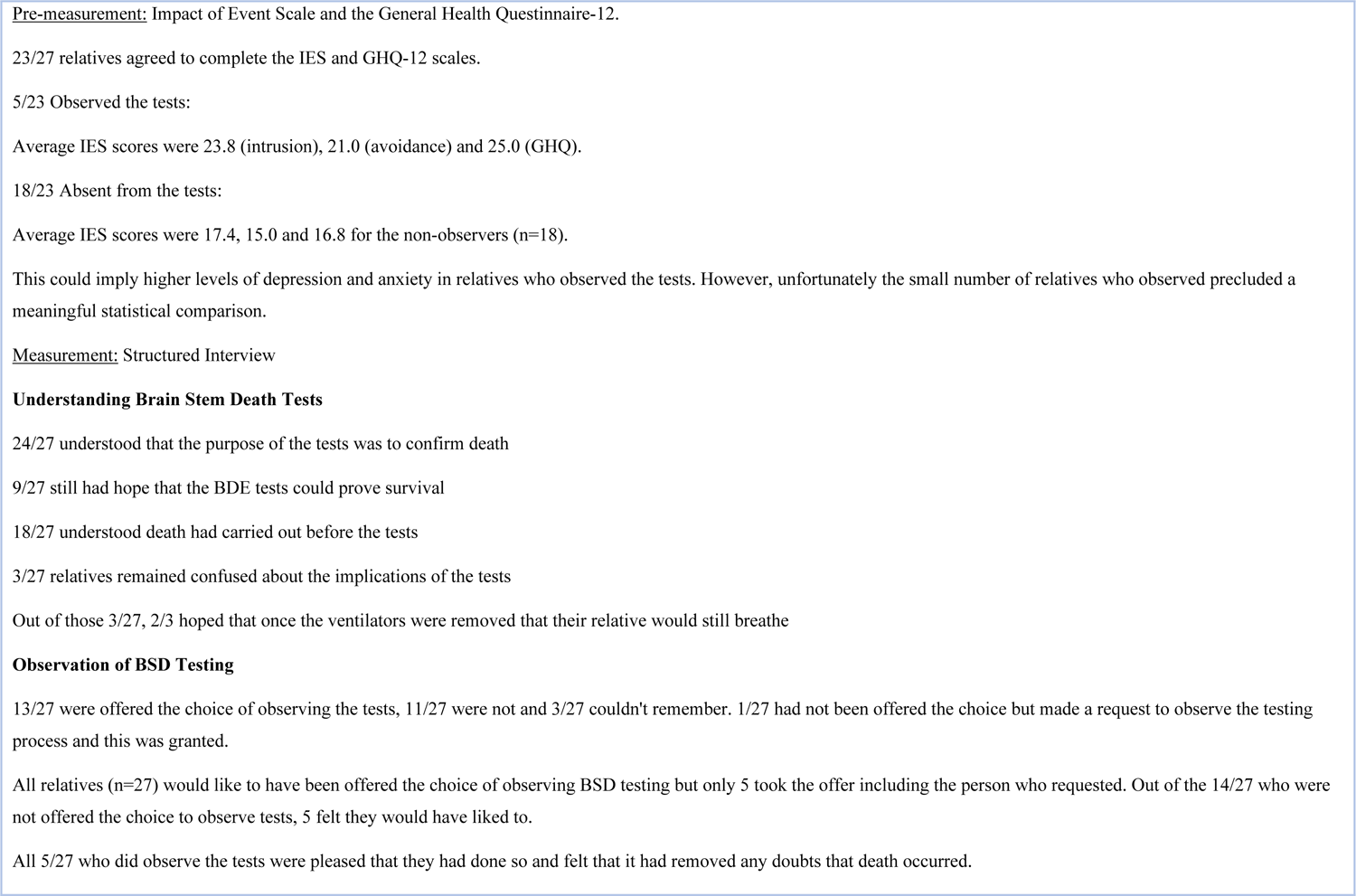
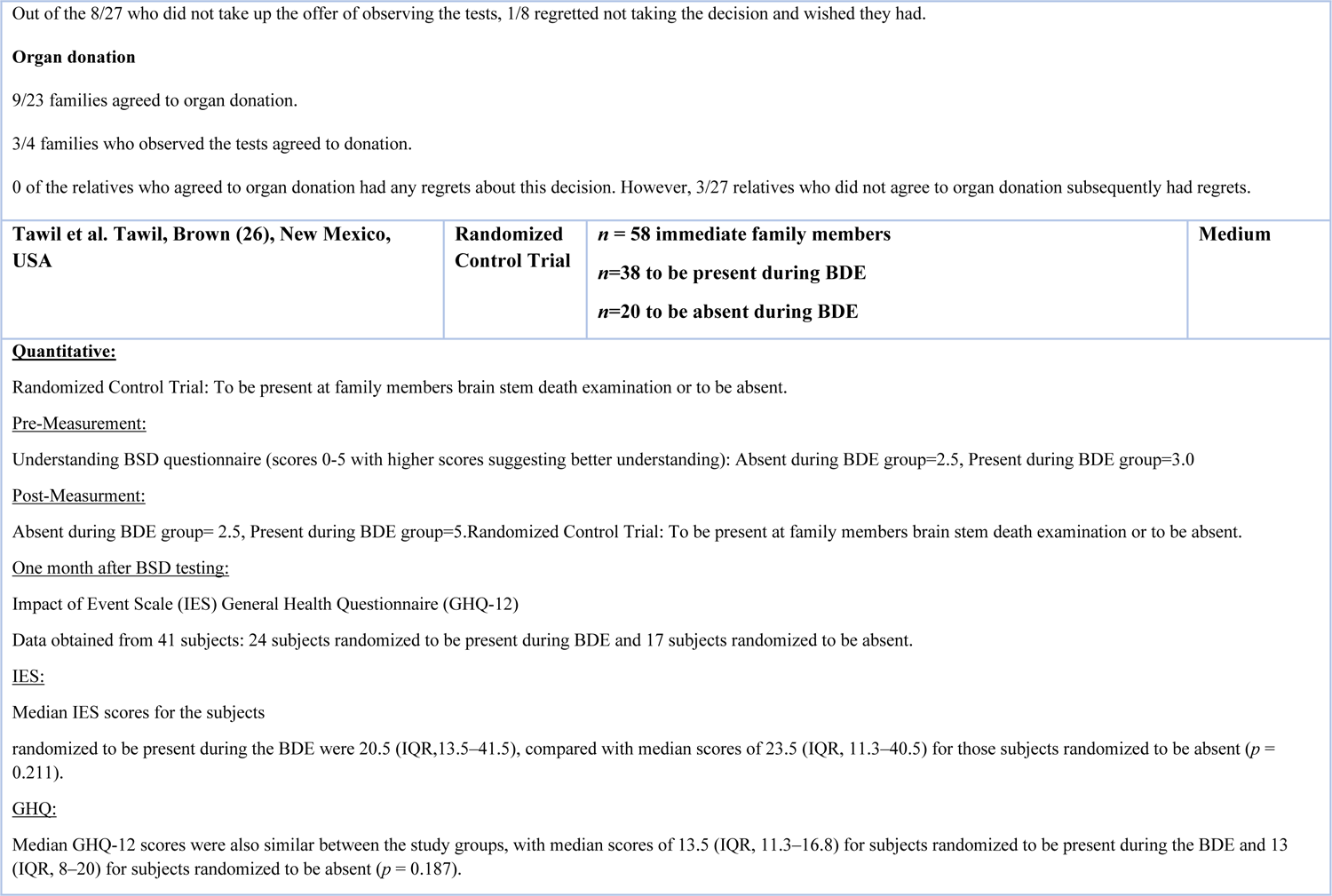
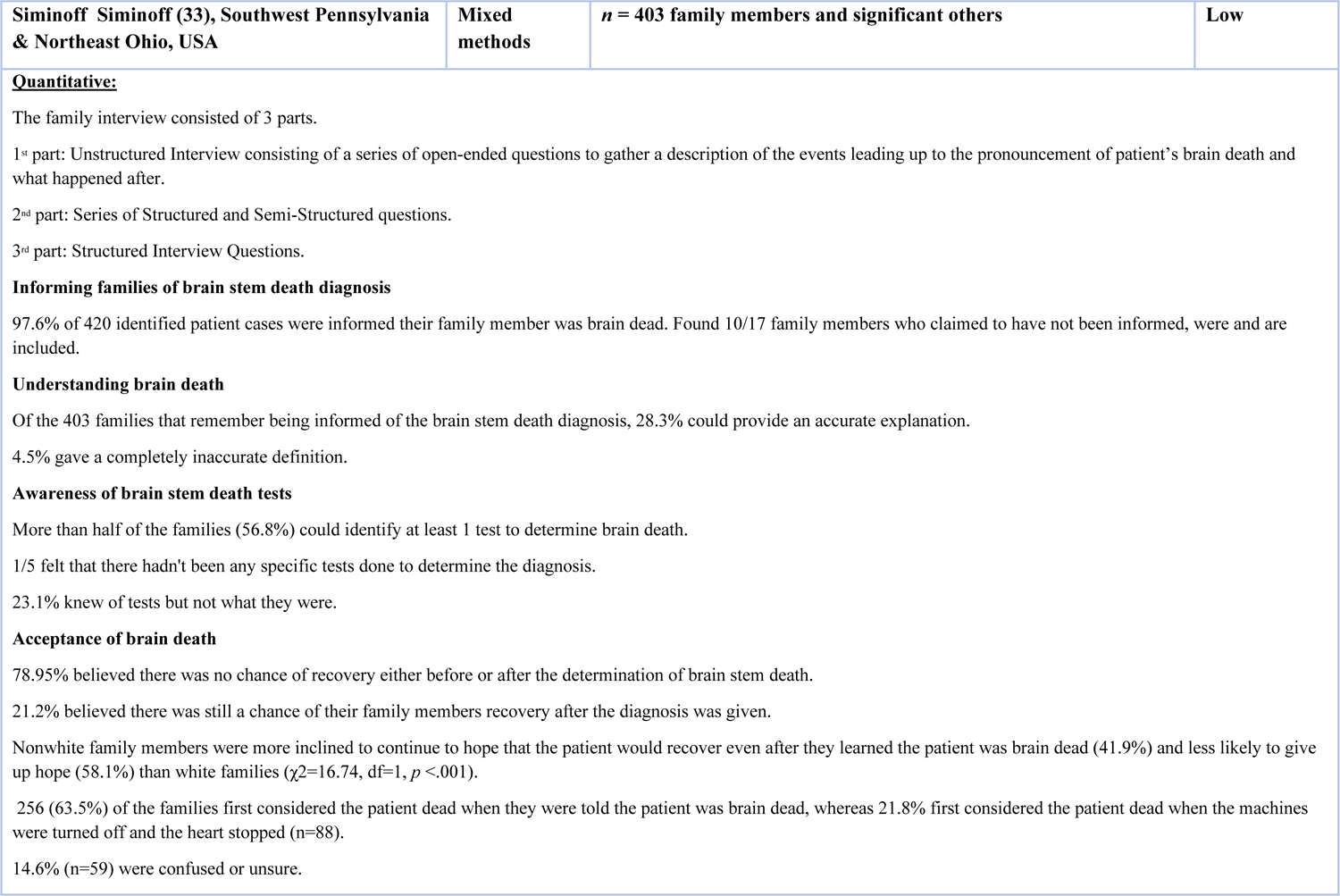
Summary of Research Papers Included in Review

### Quality assessment

Most studies were rated as being low quality (i.e., high risk of bias; *n* = 4), whilst three studies were rated as being of medium quality and two as high (i.e. low risk of bias). Most qualitative studies failed to consider the influence of the researcher on the research as authors did not locate themselves theoretically and/or culturally and there was not congruence between the research methodology and the methods used to collect data (e.g., using grounded theory but failing to use an iterative process in terms of participant recruitment).

The RCT by Tawil et al. Tawil, Brown (26) was scored low because it was unclear whether the treatment groups were similar at baseline and whether the outcome assessors were blind to the treatment allocation.

Finally, the mixed methods study by Omrod et al., Ormrod, Ryder (27) scored low because the different components of the study were not effectively integrated to answer the research question.

### Narrative Synthesis

Results were synthesised based on similarity across the included studies, placed in chronological order and relabelled. This resulted in six overarching themes (Table 2).

**Table 2.** Narrative synthesis: thematic map

| Main Identified Theme | Sub-theme |
| --- | --- |
| 1. The unexpected prognosis |  |
| 2. Coming to terms with Brain Stem Death- grieving process: | ○ Denial: Lack of understanding and unanswered questions<br>○ Bargaining: Understanding but not believing and the hope for survival<br>○ Acceptance |
| 3. Request for organ donation: | ○ The impact of acceptance<br>○ Providing a purpose<br>○ An unwanted responsibility and cause for additional distress |
| 4. Observing Brain Stem Death testing: | ○ An uncertain process<br>○ An inconsistent offer<br>○ Increasing understanding and acceptance |
| 5. The effect of staff on family's experience: | ○ Feeling seen by staff<br>○ Feeling neglected by staff |
| 6. The lasting impact: | ○ Experiencing grief<br>○ Impact on well-being |

## 1. The unexpected prognosis

Families experienced difficulties with the suddenness in which they received news of their family members’ imminent or confirmed BSD diagnosis.

Frid et al. Frid, Bergbom (28) talked about the shock participants felt at the beginning. They identified “The disquieting event” including the sudden change, the growing seriousness, and the great chaos.

Lloyd-Williams et al. Lloyd-Williams, Morton (32) also referenced experience of a sudden change, the event happening too quickly for the relative to understand; “*Her death was so sudden, so quick. The doctor said, ‘You know your wife is going to die’. I was staggered. In a matter of hours, she went from completely fine to being told that. I was totally gob-smacked. All too sudden. She was such an alive person”*.

Frid et al. Frid, Haljamae (29) reported imagery used to describe participants first encounter with the BSD prognosis that included an empty room, murder, the collapse of life and descending into hell. Kesselring et al. Kesselring, Kainz (31) identified family members’ travelling to the hospital, ‘often in a state of shock’ after having witnessed the catastrophic event themselves or having received the news from services or friends. They found the family member would then be informed immediately or hours later that they should expect ‘bad news’. This ‘bad news’ would be delivered privately or in public waiting rooms, by professionals they knew or those that they did not. This uncertainty added to the chaos they were experiencing, the disintegration of the life they once knew. Kentish-Barnes et al. Kentish-Barnes, Cohen-Solal (30) also report a feeling of being in another world, or as though the world that was known to the participant is ‘collapsing’.

## 2. Coming to terms with Brain Stem Death

This theme has been split into 3 stages (denial, bargaining and acceptance) that resemble some stages of Kübler-Ross Kübler-Ross (34) staged grieving process. Denial, which includes a lack of understanding and unanswered questions, bargaining, in which there is understanding but not believing and having the hope for survival, and acceptance.

### Denial: Lack of understanding and unanswered questions

In each study, there was a focus on family members understanding of their relatives’ brain stem death diagnosis. In Siminoff’s Siminoff (33) study, out of the 403 family members informed of the brain stem death diagnosis, only 28.3% could provide an accurate explanation of the diagnosis and 4.5% provided an entirely inaccurate explanation. Kesselring et al. Kesselring, Kainz (31) found that none of the participants they interviewed could reproduce a ‘scientific’ description of BSD. For many, it was difficult to differentiate between BSD and states like a coma, despite being given verbal explanations and documents.

For some, even witnessing the BSD testing left them with a confused understanding. Kompanje et al. Kompanje, de Groot (18) revealed that many did not realise what the tests were supposed to demonstrate, or what they would entail. Some had thought the tests could prove the potential for survival, and not as a way of confirming the patients BSD which demonstrates a lack of communication from their relatives’ physicians (27). Ormrod et al. Ormrod, Ryder (27) found some similar results, 9/27 family members still had some hope that the tests would demonstrate potential survival, 3/27 remained confused about the status of death after observing the tests, and 2 out of those 3 hoped that even after the ventilation was withdrawn, the BSD patient might still breathe.

### Bargaining: Understanding but not believing and the hope for survival

Some people when interviewed demonstrated a true understanding of BSD and what the implications were. However, that understanding did not stop the families hope for survival, even if they knew it was not possible. This conflict was often referenced because of the body being warm, and the ventilator still causing the body to breathe and appear alive.

The participants in Kentish-Barnes et al. Kentish-Barnes, Cohen-Solal (30) appeared to imply that though they had understanding, to feel fully convinced they needed to see the body after the ventilators were turned off. The warmth of the body and movement of the chest whilst the ventilators caused the patient to ‘breathe’ would seemingly lead families to experience an inner conflict; a clinical understanding of their family members’ reality whilst being confronted with the experience of their loved one appearing alive; *“We kept on touching her, all the time, she was warm….so she was there. And frankly, when I think about it now, I know that during all that time we believed that she would come back. Even if we knew it wasn’t possible”*.

Others felt that they understood the concept of BSD, but due to not witnessing the BSD tests they did not feel able to accept or believe it. The BSD testing was believed to provide a finality for some that they were unable to have if not allowed to observe (27).

Siminoff’s Siminoff (33) study found most families accepted that their family member would not recover either before or immediately after the pronouncement of brain death (78.95%), but 21.1% still felt that their relative may survive and recover. Despite this, over half (60%) of families made statements indicating they believed the patient was still alive after being told their family member was brain dead (*n* = 241). It was found that those who still believed were generally older than those who fully understood that the relative had passed away (Mean age = 45.61 vs 42.83 years).

### Acceptance

Acceptance of a situation is to recognize, endure and believe something to be true. The experience of acceptance will exist at different levels and develop over time.

Frid et al. Frid, Bergbom (28) found that for some family members, acceptance occurred once they were able to realise their loved one had passed despite them showing signs of life (breathing, warm skin etc). Kesselring et al. Kesselring, Kainz (31) also found some families could accept their relative’s death despite showing signs of life. However, they also found that others needed to see the relative once the organ donation operation was complete. In this state, the body would be cold and unmoving, having most likely been in the morgue. This enabled the families to witness their family member in a condition that fit their pre-existing understanding of death, rather than warm and seemingly breathing; *“After the operation, I was able to see him once more. I was very happy and seeing him cold was good since you cannot say goodbye when he is still breathing”*

For some, observing the BSD testing enabled the family member to have closure. Ormrod et al. Ormrod, Ryder (27) found that all five relatives who observed the tests were pleased that they had done so and felt that, for them, it had removed any doubts that death had occurred.

Siminoff Siminoff (33) referred to the concept of acceptance as “Giving Up Hope” which was related to the family’s ethnicity. Non-white family members were less likely to give up hope (58.1%) than white families and more inclined to continue to hope that their family member would recover even after learning the patient was brain dead (41.9%). Furthermore, 63.5% of the families first considered their relative to be dead when the announcement of the brain death diagnosis was given, whereas 21.8% first considered their relative to be dead when the machines had turned off and their heart had stopped. Moreover, they found that those with lower-level education were more likely to agree with the statement that a person is only dead when their heart stops, and that non-white family members were more likely to agree with this statement than the white family members.

## 3. Request for organ donation

### Impact of acceptance

Siminoff Siminoff (33) found that the families understanding of brain stem death was not an indicator of organ donation, as demonstrated by the fact that 60.2% of the people who said that brain dead individuals were still alive chose to donate. However, acceptance of the family members death did correlate with families organ donation decision. Firstly, the families who accepted that their loved one had died when told that they were brain dead were more likely to donate than those who thought their loved one had died once the machines had been turned off and their hearts had stopped (62.5% vs 39.8%). Secondly, those family members who were ambiguous as to exactly when their relative had died had a similar pattern of donating to those who understood brain death to be death (62.7% vs 62.5%).

Moreover, it was found that those who agreed to donate their relatives’ organs were less likely to agree with the statement that someone is only dead when their heart stops than those who did not donate.

### Providing a purpose

Frid et al. Frid, Bergbom (28) found that for those who had a good idea of what their family members would have wanted, making the decision of organ donation seemed like a way of connecting with them.

Frid et al. Frid, Haljamae (29) found the imagery that came up was that of a mutilated body. The participant felt that though the organ donation process may involve a ‘violation’ of the deceased, it is the ‘right thing to do’ if it can save someone’s life. Some of the participants in Kentish-Barnes et al. Kentish-Barnes, Cohen-Solal (30) study expressed this and the strength it provided them in a time of grieving, as well as a belief that it led to a continuation of the relatives’ memory.

Ormrod et al. Ormrod, Ryder (27) found that 9/23 of the families agreed to organ donation, none of which reported to have regretted this decision afterwards. Out of the 4 families who did observe the BSD tests (5 relatives in total), 3/4 agreed to donation. Furthermore, Ormrod et al. interviewed patients who had received contact from the transplant recipients, which provided comfort and a feeling of connection with the families who had been positively impacted by their relative Ormrod, Ryder (27). Organ donation can also provide a space for grieving and acceptance in families’ experience of BSD. For some, the time and process around organ donation provided a space for reflection in an otherwise bewildering situation (30); *“So I used that time that was actually for the organ donation process, I used it to accept a situation that was not even conceivable a few hours earlier there. So that was extraordinary in fact”*.

### Unwanted decision and cause for additional distress

Though for some organ donation can be a positive experience, for others it can provide additional distress in an already very painful situation.

Frid et al. Frid, Haljamae (29) highlighted two notions people can experience; the ability to see the person as indivisible and divisible. For some, the person is a whole body, a unit, in which case the concept of organ donation is very difficult.

Another point of distress can come from the family members having to weigh up their own views on organ donation, and those of the deceased (30). If these do not align, the process can be very conflicting. Furthermore, some families do not feel it is their place to make the decision at all (30).

Kesselring et al. Kesselring, Kainz (31) found some family members’ reference witnessing the donor management, finding watching the disturbances that were made to the body led to ambivalence surrounding their decision; *“They took lots of blood, made x-rays and so on and you start to doubt whether all these disturbances to the body are really necessary? … What have I done?”*.

In addition, others (due to being asked regarding organ donation when they were still not fully accepting or understanding of BSD) spoke of finding the process especially distressing, wondering afterwards whether they had “killed (them) by consenting”.

However, Ormrod et al. Ormrod, Ryder (27) found no difference in recorded IES and GHQ-12 scales in those who did donate and those who did not donate their organs. 3 relatives who denied consent for organ donation in their study reported regretting their decision, and some felt that their original decision was linked to them having not observed brain stem death testing, perhaps highlighting the potential influence of observing BSD tests on organ donation.

## 4. Observation of Brain Stem Death testing

### An uncertain purpose

The results highlighted the lack of information that was provided by professionals about the tests, both in terms of the process and what the individual tests entail. This appeared to add to the family members uncertainty surrounding an already overwhelming situation.

Ormrod et al. Ormrod, Ryder (27) studied the impact of families observing the BSD tests and found that 24/27 participants understood that the purpose of the tests was to confirm BSD. However, 33% still had hope that the tests could prove potential survival had occurred before they were carried out. 11% (3/27) were confused about the implications of the tests, with some (2/3) people hoping that even after the ventilators were withdrawn, their relative may still breathe alone and survive.

Siminoff Siminoff (33) found that more than half of their participants could identify at least one test to determine BD. However, 23.1% knew of specific tests used but could not provide any explanation or if they did, they spoke of misconceptions. 1/5 even felt that there had not been any specific tests done to determine the diagnosis of their loved one.

This uncertainty was also cause for additional distress for family members. Participants stated concern that the tests would inflict pain on their relative. Some found the tests quite difficult to watch, especially if the tests were not explained to them throughout. On the other hand, if the family had not observed the BSD tests, some of their imagined ideas of those tests could also cause distress.

### An inconsistent offer

Ormrod et al. Ormrod, Ryder (27) also found that out of the 13 participants who were offered the choice of observing the tests, 11/27 claimed they were not offered this opportunity and 3/27 said that they could not remember. One of the participants who was not offered the choice, requested they see the tests which was granted. All participants included in their study *(n =* 27) said afterwards that they would have liked to have been able to observe the tests, but only 5 accepted this offer, including the participant who requested. Out of the 8 participants who were offered the opportunity to observe but declined, 1/8 regretted not making the decision and wished that they had.

### Increasing understanding and acceptance

Tawil et al. Tawil, Brown (26) conducted a randomized control trial and found that observing the tests led to an increased understanding of BSD. They measured family members understanding of BSD prior to randomizing them into two groups, one of which would allow the family members to be present whilst the BSD tests took place, and the other where they were absent. At a baseline, the group which would be present had a median score of 3/4 and the group who were not going to be present in the BSD tests had a median score of 2.5/4. However, when the groups understanding was measured immediately after the experience, those who were randomized to be present for the BSD tests had increased scores and therefore increased understanding of brain death, whereas those who were randomized to be absent from the tests demonstrated no change in their understanding. Furthermore, 66% of those who were present during the BSD tests achieved perfect post-intervention BSD understanding scores, compared to only 20% who were absent (median 5.0 vs 2.5).

In addition, their study demonstrated that there was a correlation between those with a higher education, who were earning over $25,000, who were younger and those who understood and could describe BSD tests. 36/38 of those who were present during the tests reported that being able to observe them had helped their understanding of BSD. In addition, 32/38 said they would recommend family presence during the tests to others going through the same experience.

In addition, Ormrod et al. Ormrod, Ryder (27) found that out of the 14 who had not been offered the opportunity to observe the tests, 5 said they would have liked to in retrospect. All 5 participants who did observe the BSD tests were pleased they had done and reported that it had removed any doubt that their family member had died. For some, seeing the tests only confirmed what they already knew, and was seen as more of a formality.

Moreover, other interviewees reflected on the potential usefulness in observing BSD testing for decision making surrounding organ donation, wondering whether perhaps the tests would have provided confirmation of death, resulting in making the decision process easier.

## 5. The effect of staff on family’s experience

### Feeling seen by staff

Encounters with the carers are perceived as important to families, demonstration of empathy and genuine sympathy are key in making the families feel supported and ‘seen’ (28).

Having both the family members and the BSD relative treated with respect and cared for was important for a positive experience. This included communication regarding the BSD, taking initiative to answer any unanswered questions, treating the families loved one with dignity (like being washed) and communication to the loved one at times too (28, 31, 32); *“They saw if we looked questioning at them and they provided information of their own accord. One didn’t need to ask, that I thought was really good.”* (28)

Specific support for organ donation surrounding any misunderstanding or religious needs were also provided, leaving families feeling supported and heard (31); *“[The patient’s] greatest fear was organ trade, he felt very strongly about it. Two physicians took their time, sat with us 2.5 hours and really explained everything to us … They gave us time to discuss things and offered psychological support if we needed it. It went very smoothly”*

### Feeling neglected by staff

In some papers, staff were reported to have been ‘too busy’ to spend quality time with families to make sure they were ok and understood what was happening (28). Family members wished they had received more support and guidance from those in the know (32).

Another issue that families confronted was a lack of communication or miscommunication which led to distress, confusion, and unanswered questions (28, 31, 32); *“Nobody said anything to us there – nobody communicated with us. Nobody came out and talked to us during those days – we were told absolutely nothing.”* (28).

Families not feeling heard or respected often centred around organ donation, with families refusing donation and being asked again afterwards (31).

## 6. The lasting impact

### Experiencing grief

Family members can often still have unanswered questions about BSD, long after the experience is over (28). Further to this, with the BSD diagnosis and organ donation experience happening so quickly, the last image families can have of their family member is of them warm and breathing (due to the ventilator). This can cause people to feel as though they are saying goodbye to their loved one whilst they are still alive (30). Others did not feel they were able to say goodbye to their family members at all because of the difficult ending as well as not being informed where the patient would be taken to (28).

Moreover, observing BSD testing without a full explanation of their purpose and what they entail can potentially leave families with more distress (27). Though, for those who did not observe the BSD testing, not knowing what the procedure included led them to have negative images of what they imagined it to be (27).

A way people were able to cope is by finding meaning in what happened. For example, knowing that if they had survived, they may not have had a good quality of life (28). However, many families reported wanting further support after their experience in ICU, feeling it would have helped them deal with their grief (28, 30, 32); *“A letter after the event, saying something like, ‘We understand how things are. In the meantime, here is a number.’ You are on your own afterwards. You have got time to think. It would be nice if they would see if you would like to attend a support group or answer questions, to know that you are not on your own. It would be good if they let you know straight away.”*(32).

### Impact on well-being

Tawil et al. Tawil, Brown (26) and Ormrod et al. Ormrod, Ryder (27) measured participant well-being utilizing the Impact of Event Scale (IES) and General Health Questionnaire-12 (GHQ-12).

Tawil et al. Tawil, Brown (26) obtained 41 subjects post measurements (71%): 24 subjects which were randomized to be present for the BSD tests and 17 who were randomized to be absent. The IES and GHQ-12 were similar for both groups. The median IES results for those who were present for the tests was 20.5 and the results of those who were absent were 23.5. In addition, the GHQ-12 scores were also similar between the groups. The participants in the group who were present in the tests had a median score of 13 and those who were absent had a median score of 13.6.

Ormrod et al. Ormrod, Ryder (27) found that the average IES scores of the 5 relatives who observed the tests were 21.0 (avoidance) and 23.8 (intrusion) and the average GHQ-12 score was 25. This was compared to the IES & GHQ-12 scores of the rest of the participants which was 15 (avoidance), 17.4 (intrusion) and 16.8 (GHQ-12). These results could suggest that there were higher levels of depression and anxiety in those who observed the tests, though due to the small number of relatives who observed the tests it is difficult to create a meaningful statistical comparison.

## Discussion

The narrative synthesis highlighted six key common themes across the included studies: The unexpected prognosis, coming to terms with brain death-grieving process, request for organ donation, observation of brain stem death testing, impact of staff on family’s experience and the lasting impact.

One of the main findings is that families often do not understand the concept of BSD, what the diagnosis is, and what the implications are. Research suggests that patients only accurately recall around 49% of information communicated to them by healthcare professionals when prompted (35) and that 40-80% of information given by healthcare professionals is often forgotten immediately (36). This highlights the importance of giving information in multiple formats as verbal comprehension and recall can be compromised in distress.

A lack of communication and input from staff forces families to have to work out the answers for themselves and considering the wide range of debate surrounding BSD online, time in the ICU with expert professionals presents an opportunity for clarification that families may not receive elsewhere. Research has shown that families in ICU often perceive staff as being busy and unavailable (37), report having to constantly seek out staff to get updated information, being rarely approached by staff to ask if there was anything they needed to know and often not provided with a private space to discuss news with professionals (38). The findings highlight the importance of staff input on family’s experience and need for better family support provision and training for clinical staff.

The review also highlighted the resulting difference between people understanding and accepting a relatives BSD. For some, despite understanding the diagnosis and the implications, their hope for their loved one surviving was not waivered. For many, seeing their relatives seemingly alive and breathing gave cause for additional distress and uncertainty. In some instances, observing BSD testing could be helpful in peoples’ movement towards acceptance of death. Many people were pleased to have been present for the testing and to have had the choice and said they would recommend it to future families. With proper communication and explanation from staff, the process was found to provide further explanation of BSD and cement peoples pre-existing understanding. However, without clarification of its purpose and what it would entail, it left some family members further confused. Hodgkinson and liams Hodgkinson and liams (39) found that with sudden death of relatives, viewing the body may increase levels of anxiety and distress in the short term, but it lessens the distress for people in the long term. Though there is little research into the effects of observing BSD tests on family’s experience, long term impact, their understanding, acceptance and grief, nevertheless, these results suggest the importance of integrating the choice for families to observe the tests.

Religion and spirituality are likely to play a role in this experience of holding on to hope vs acceptance, like in the case of Jahi McMath (40). The concept of brain death is less than 100 years old and therefore religions and their many denominations may have various beliefs on whether brain stem death is believed to be ‘death’ (41). This highlights the importance of involving Chaplaincy or equivalent services in supporting families. Many believe that grief plays a particularly poignant role in the ethical encounter of brain death as it begs the question of what constitutes life and death (42). We can currently recognise grief through a variety of psychological models. Often, they appear linear as if the journey of grief has an end. However, the Kübler-Ross Kübler-Ross (34) model looks at 5 stages of grief that one can experience in any order. Kubler-Ross based this understanding on her work within palliative care, the stages include; denial, anger, bargaining, depression and acceptance. In many models of grief, the idea of ‘acceptance’ is key.

The allowed time for ‘acceptance’ and understanding of the diagnosis is referred to in BSD literature. In 2014, the Neurocritical Care Society (43) published a toolkit online regarding brain death to help hospitals in the US modernize their policies around the determination of BSD. When considering the communication with family, they refer to a ‘reasonable amount of time’ being allowed for the family to see the relative and come to terms with the diagnosis before the ventilator is removed. This implies that grief can be ‘handled’ by allowing family to spend some time with their family member so that they can come to accept the bereavement Friedrich (42).

Organ donation as a process was reported to have provided some additional time for the family to process their experience as well as reassurance, knowing the tragedy could have some purpose in saving another’s life. Although this seemingly depended on the level of communication received. Often families did not feel as if they were fully informed of the organ donation procedure, nor were they granted the time to come to terms with the events. Being asked to donate family members organs without either fully understanding the implications of BSD or without having ‘given up hope’ could lead to further distress (31). It left some to wonder whether their own decisions had contributed to their loved one’s death. Siminoff Siminoff (33) found that understanding of BSD did not correlate with the rate of which people donated, in fact they found that some who did not understand or accept their loved ones’ death still agreed to donation.

Other studies focusing on organ donation have found that families require simple, clear, accessible information about the cause and diagnosis of BSD and decision making in small bitesize pieces, including this time to think and ask questions to best support people and encourage donation (44). Some participants also described it as a *‘suspended death’* (28) describing a discrepancy between what they felt was the time of death and the medicolegal death which they claimed interfered with their grieving process (death anniversaries unknown, for example). Boss Boss (45) identified a new type of loss named ‘Ambiguous Loss’. It occurs when a loved one is physically present but not psychologically, like Dementia. Boss states that because the loved one is here, but seemingly not, grief can be frozen or ‘put on hold’, which leaves people traumatized (46). It is important to consider the idea of ambiguous loss when thinking about BSD, the family member appearing warm and breathing when told their life is permanently lost.

When considering families well-being scores, research included within this review has mixed results. Tawil et al. Tawil, Brown (26) found no significant difference in family members’ median IES and GHQ-12 scores of those who observed brain stem tests and those who did not. However, a moderate or severe impact of trauma requires a score of over 26, with 33 and above being considered as the best representation for a probable PTSD diagnosis. Both groups, present and absent, reported their highest IES to be above 26. In addition, though no significant difference was found between the two groups’ median, the group which was present for the BSD testing was found to have the highest scores, both reaching above the score of 33 (avoidance and intrusion scores). Furthermore, Ormrod et al. Ormrod, Ryder (27) found in their study that observing the tests led to an increase in the likelihood of depression and anxiety. Findings highlight the need for long-term psychological support for families.

### Strengths and Limitations

This review is the first to synthesise families experience of a relatives brain stem death diagnosis and highlight potential clinical and research opportunities that can better support families. The quality assessment and full text screening was carried out independently by two authors to reduce the level of bias.

It is also important to consider the strengths and limitations of the topic area. The review consisted of 606 participants overall *(n =* 403 from one study), thus additional research would need to be produced on this topic to ensure a larger sample could be reviewed across a range of settings. In addition, only 3 studies with quantitative data were included and therefore the heterogeneity in study methodology and populations limited a meta-analysis. Furthermore, the quality assessment of the data showed 4/9 studies included had a high risk of bias and low publication quality indicating the need for higher quality research.

The diversity of practice could also be considered a barrier in conducting this review. The wide range of variables including healthcare settings (private, public, understaffed etc), specific country or state guidelines surrounding BSD, mandatory protocols and tests used to identify BSD are all important variables to consider that will impact a family’s experience which often were not specifically identified within the individual papers themselves.

Moreover, very few studies focused on the cultural impact of both researchers and participants when experiencing BSD, especially given the role of spirituality and religion in the experience of understanding, and accepting death. Quality assessment highlighted that cultural or geographical location was not considered when examining peoples experience of BSD, demonstrated through a lack of statement from the authors locating themselves and the lack of demographic information included, with only 2 referencing ethnicities.

### Implications

#### Intensive Care Unit Support Protocols

Though countries may differ on their diagnostic criteria for BSD, there is a possibility for a unified perception or method of practice when involving and supporting the families.

Initial care families receive has an impact on grief and that the way families are initially informed of their relatives BSD has a significant impact on their overall understanding (47). Not only does the explanation of BSD need to be uncomplicated and accessible, but that support is provided throughout each stage and in different formats (verbal and written) so that the explanation can be reiterated to confirm understanding and staff are present for any further questions (48, 49)

Caswell Caswell G (50) conducted a study of communication between families and staff in end-of-life care. They found that staff could recognise the desire to have more than one conversation with relatives but also acknowledged that they were often reluctant to want to talk about death. Reportedly in most countries, how to speak with patients about death is not included in medical school and allied health professional’s education (51). Therefore, it is important that professionals have a protocol they can follow to allow for adequate support, consistency in communication and confidence in providing care under these incredibly sensitive circumstances. If meaningful staff involvement is facilitated via implemented care protocols within these settings, family members’ psychological needs can be met and their grieving process supported (52).

These protocols could include; private locations in which families can be spoken to regarding the BSD diagnosis and what it entails, psychoeducation, a uniform description that professionals within ICU are familiar with and feel confident in talking through to avoid misinformation or inappropriate language, having an allotted case worker/staff member to whom any questions or concerns can be raised to help the family throughout the process, allowing families time with their relative to give space for acceptance, working with the families to find out what support may be best for them in the future and arranged check-in calls to follow up on any potential mental health support needs and signposting services/resources to support with their grief.

The John Hopkins Hospital, USA created its first Death by Neurological Criteria Team in June 2016. This team’s development means that diagnosis of BSD can happen faster as a result of specific professionals who are experienced in BSD working together to conduct the tests and support the families (53).It is of interest to see the evaluation of such services and how they benefit families.

Given the families’ unique experience of grief, and the apparent confusion and uncertainty surrounding official announcement of BSD, families in this situation require specific grief support to help navigate them throughout this incredibly difficult time. Volpe Volpe (54) argued that in order to care for the BSD relatives’ families effectively and manage the inevitable ethical and moral conflicts surrounding the diagnosis, there needs to be further research into new understandings of grief and ways in which that understanding can be incorporated into the communication with these families.

#### Trauma-informed support

Kristensen Kristensen (55) argues that support for those family members who have experienced sudden and violent deaths of loved ones may require different grief support interventions than for loss from natural death. This is because several studies have shown that these sudden bereavements can adversely affect mental health of close ones, with mental health disorders appearing more elevated and with recovery taking longer. Studies also show that peoples initial reactions to the news of sudden death (like denial or guilt for example) can assist professionals in estimating how long or severe the process and progression of grief may be, whether traumatic or pathological (56, 57). This early identification of traumatic grief can enable appropriate follow up and specific support.

Bolton Bolton JM (58) found that those who grieve after unexpected death are three times more likely to have psychological disorders than those who grieve a natural death. Though the reviewed studies did not explore any potential trauma symptoms associated with the experience of their loved one passing, Tawil et al. Tawil, Brown (26) demonstrated how this experience could potentially lead to symptoms of PTSD and some participants referenced specific distress surrounding imagery and memories from their hospital experience. For example, though for some witnessing the BSD tests helped with their acceptance of the loved ones diagnosis, some referenced experiencing intrusive memories over a year later and even those who did not witness the tests found the imagined version of them seemingly traumatic,; *‘The most vivid parts of the tests was the cotton wool – I don’t know why but I remember it over and over again.’;‘I visualise the tests even though I never saw them”.* (27). Scott Scott (59) found that social support following a sudden bereavement is associated with reduced depressive and PTSD symptoms. Thus, further research into trauma-informed grief support needs of the family members affected both in the short and long term are needed.

#### Cultural Implications

As shown in Siminoff Siminoff (33) cultural differences can impact the way in which we understand death and are able to accept it. It is important for us to examine the way in which this not only will impact families experience of BSD, but also the way in which professionals may understand it too. Consequently, additional implications to consider could include providing a safe space for professionals themselves to ask questions. It is important to have a workforce which is knowledgeable and respectful, understanding not only the different beliefs that could exist between their service users, but also within their team, encouraging discussion without judgement to ensure their well-being is also considered. If physicians are well-informed in this way, they can then better manage sensitive discussions with families and reduce confusion (14).

Moreover, working closely with chaplaincy services and religious leaders can increase trust of the medical interventions and help families feel at ease (19).

## Conclusion

This review demonstrates the psychological impact of a relatives’ brain stem death diagnosis on families, most notably families lack of understanding of the brain stem death diagnosis, even months after their relative has passed and longer-term psychological distress. It is of importance to establish specific BSD protocols that can aid a consistent approach, better communication with families and further research into ways in which families can be supported throughout this process to avoid adverse consequences and psychological distress.

## Data Availability

All relevant data are within the manuscript and its Supporting Information files.

